# Doctor-Patient Communication and Sensory Disabilities among Community-Dwelling Older Adults: Findings from NHATS

**DOI:** 10.1101/2025.07.22.25332001

**Authors:** Kevin E. Nguyen, Theodora Papachristou, Olga Korosteleva

## Abstract

**Objectives:** Older adults with communication disabilities (CDs) encounter challenges across various environments, with the most pronounced difficulties arising in healthcare settings. While CD is a common occurrence, limited research explores the medical and socioeconomic factors contributing to CD in the patient-centered setting. Therefore, our paper explores this interplay between communication experiences for older adults in healthcare contexts.

**Methods:** We combined and cleaned data (N=3725) from 2011 (Round 1) to 2022 (Round 12) taken from The National Health and Aging Trends Study (NHATS) and employed a binary logistic model and a weighted model to account for population adjustment to identify prevalent covariates influencing our hypothesis.

**Results:** In a sample of 3725, health-related concerns (OR = 11.06, 95% CI: 7.81–15.65) significantly contributed to communication difficulties in the doctor’s office. Hearing loss (OR = 1.56, 95% CI: 1.05–1.10) and low education levels (below high school) (OR = 1.59, 95% CI: 1.23–2.06) were also associated with communication challenges.

**Conclusion:** These findings indicate that significant health issues in adults may contribute to the development of communication disorders, underscoring the need for comprehensive support mechanisms. Future research should focus into supportive strategies and improved physician relationships is vital to support the needs to disabled patients.

## Introduction

Approximately 15% of American adults, making up a population of 37.5 million, reported experiencing some degree of hearing difficulty. This number rises sharply with age, as about one in three people in the U.S. between the ages of 65 and 74 encountered hearing loss. By the age of 75, nearly half the adult population experience hearing difficulties, leading to problematic communication in medical settings. In such settings, Effective communication is critical to physicians to ensure a nurturing patient-centered atmosphere [10]. Ideally, effective doctor-patient communication ensures a reciprocal process of decision making in which the opinions and wishes of patients regarding their health care are respected and considered. This process may pose significant challenges for older adults who may experience unique communication barriers such as having someone sit in at the doctor’s visit to articulate the older adult’s health concerns, explain treatments or remind the patient of questions to ask. To ensure proper support for at-risk patients, accompanied beneficiaries whose visit companions were more actively engaged in communication rated physician information giving (OR, 1.42) and interpersonal skills (OR, 1.29) more favorably.” We believe these results, taken together, merit the attention of policy makers, physicians, and researchers interested in improving quality of care for the sickest patients, who tend to be least satisfied with the quality of their health care [14] Hearing loss, a prevalent condition among aging adults, rises with advancing age. Individuals with hearing loss visit physicians significantly more than those without such conditions, yet may be underutilizing available services [20]. Since hearing loss is a significant issue, higher rates of hospitalization and complex interactions with healthcare providers remain. Such loss impedes effective communication during clinical encounters, putting patients’ safety at risk, leading to delayed diagnosis, improper treatment decisions, and reduced adherence to care plans.[8]

Qualitative research linked effective doctor-patient communication to benefits, such as improved health outcomes [9], improved adherence to medication and treatment [6], overall improved patient satisfaction [5,14,21] and trust [3] towards their medical provider. Yet, quantitative research on factors influencing effective doctor-patient communication is bounded by differing measures, differing settings, i.e. hospital versus doctor office visits, differing methods of data collection, i.e., self-reports versus clinical measures and differing populations.

In our study, we are examining characteristics that may contribute to older-aged patients experiencing communication difficulties (CD) experienced with the doctor in the medical room. Previous research hints underrepresented minorities (such as Blacks) as having a greater risk of underutilizing disability support. Additionally, there is limited research on the utilization of support individuals during medical visit [15]. To fill in this gap, we aimed to investigate the main factors such as demographics, chronic health conditions, and functional statuses, that lead older adults to face challenges in doctor patient communication in the medical office. Our research examines this relationship at two levels: at the sample level and adjusted to the population level. The remainder of the article delves into the underlying factors affecting participants’ responses to questionnaires, which are believed to influence communication disabilities. We hope that these findings will not only contribute to future research but also bridge the communication gap between doctors and patients and potentially improve medical outcomes.

## Methods

### Study Sampling Design

The National Health and Aging Trends Study (NHATS) is a comprehensive resource that explores the health and functioning of older adults in the United States. This longitudinal study focuses on Medicare beneficiaries aged 65 and older, aiming to provide valuable data on how aging affects daily life, health outcomes, and overall well-being. According to projections by the

U.S. Census Bureau, by 2035, older adults are expected to outnumber children, with an increasing risk of disability as age progresses. NHATS supports research efforts to improve the quality of life for aging populations by offering insights that inform strategies to prevent or reduce disability. NHATS began in 2011 (Round 1) and continued through 2022 (Round 12), with each round including about 8,000 participants. The study employs a stratified, multi-stage sampling method to ensure the representation of key subpopulations, such as Black individuals and those aged 90 or older. Participants were selected based on household Medicare enrollment data, and new participants were regularly added to account for aging into Medicare, non-responsiveness, and participant mortality.

Through annual in-person interviews, NHATS gathers comprehensive data on participants’ daily activities (ADLs), economic circumstances, social mobility, health status, and more. This long-term design allows researchers to track how individuals’ health and life circumstances evolve, offering critical insights into aging. Data is collected through in-person interviews, gathering detailed information on participants’ daily activities, health status, demographics (such as educational background, income, and gender), and quality of life. The NHATS dataset is publicly accessible and can be applied across various research fields, including psychology, epidemiology, health sciences, healthcare economics, and gerontology. The valuable insights offered by this dataset contribute to a deeper understanding of aging and its impact on individuals and society.

## Data Description

### ***I.*** Data Cleaning

We began by combining datasets for sample persons (SP) who were interviewed between 2015 (Round 5) and 2022 (Round 11), creating a master dataset consisting of 12,427 participants.

From this, we refined the dataset to include only individuals who participated in all seven rounds (n=3,725). The following variables shown in Table 1 below were selected for the analysis.

**Table 1.** Descriptive statistics for analysis variables in Rounds 5 and 11.

| Variables | Round 5<br>Frequencies (%) | Round 11<br>Frequencies (%) |
| --- | --- | --- |
| **<u>Hearing/Vision Loss</u>** |  |  |
| Near Vision Loss |  |  |
| Present | 150 (4.03%) | 597 (16.03%) |
| Absent | 3575 (95.97%) | 3128 (83.97) |
| Distance Vision Loss |  |  |
| Present | 216 (5.80%) | 688 (18.47%) |
| Absent | 3509 (94.20%) | 3037 (81.53%) |
| Hearing Loss |  |  |
| Present | 842 (22.60%) | 1479 (39.70%) |
| Absent | 2883 (77.40%) | 2246 (60.30%) |
| **<u>Demographics</u>** |  |  |
| Gender |  |  |
| Male | 1531 (41.1%) | 1531 (41.1%) |
| Female | 2194 (58.9%) | 2194 (58.9%) |
| Age |  |  |
| 65-74 | 1666 (44.72%) | 468 (12.56%) |
| 75+ | 2059 (55.28%) | 3257 (87.44%) |
| Race |  |  |
| Black, Non-Hispanic | 757 (20.38%) | 757 (20.38%) |
| White, Non-Hispanic | 2685 (72.27%) | 2685 (72.27%) |
| Hispanic | 183 (4.93%) | 183 (4.93%) |
| Other Race | 90 (2.42%) | 90 (2.42%) |
| Education |  |  |
| Below High School | 663 (17.85%) | 663 (17.85%) |
| High School up to Trade School | 1224 (32.95%) | 1224 (32.95%) |
| Some College (minimum community college) or above | 1828 (49.21%) | 1828 (49.21%) |
| Marital Status |  |  |
| Married or With a Partner | 1952 (52.40%) | 1606 (43.12%) |
| Separated, Divorced, Widowed, Never Married | 1773 (47.60%) | 2119 (56.88%) |
| **<u>Medical Problems</u>** |  |  |
| Medical Management |  |  |
| Always Did it by Self | 3416 (91.70%) | 3107 (83.41%) |
| Otherwise | 309 (8.30%) | 618 (16.59%) |
| Health Condition Impairment |  |  |
| Major (Score of 3+) | 1680 (45.10%) | 2014 (54.06%) |
| Mild (Score of 1-2) | 1710 (45.91%) | 1213 (32.57%) |
| None | 335 (8.99%) | 498 (13.37) |
| Overall Health Condition |  |  |
| Poor | 717 (19.25%) | 1183 (31.76%) |
| ADL Impairment |  |  |
| Present (score of 1+) | 398 (10.68%) | 398 (10.68%) |
| Absent | 3327 (89.32%) | 3327 (89.32%) |
| IADL Impairment |  |  |
| Present (score of 1+) | 552 (14.82%) | 552 (14.82%) |
| Absent | 3173 (85.18%) | 3173 (85.18%) |
| Has a regular doctor |  |  |
| Yes | 3564 (95.68%) | 3185 (85.50%) |
| No | 161 (4.32%) | 540 (14.50%) |
| **<u>Psychological Variables</u>** |  |  |
| Rate Your Memory |  |  |
| Poor | 725 (19.46%) | 1411 (37.88%) |
| Good | 3000 (80.54%) | 2314 (62.12%) |
| Outlook on Life |  |  |
| Mild (Score 1-14) | 1069 (28.70%) | 1149 (30.85%) |
| Positive (Score 15-21) | 2656 (71.30%) | 2576 (69.16%) |

### ***II.*** Outcome Measure

The outcome variable under investigation is whether the participant encounters problems during a doctor’s visit related to vision or hearing impairments. This variable is a composite measure derived from responses to four dichotomous questions. If a participant answers "Yes" to three or more of these questions, the outcome is coded as 1 (indicating problems), while two or fewer affirmative responses result in a value of 0 (indicating no problems). The questions asked were:

i. “In the last year, did anyone sit in with you/SP and your/his/her regular doctor during your/his/her visits?”
ii. “During doctor visits within last year, did you or anyone remind SP of questions you wanted to tell the doctor?”
iii. “During doctor visits in the last year, did you or anyone ask or tell the doctor things for you or SP?”
iv. “During those visits in the last year, did you or anyone help SP understand what the doctor is saying?”

### ***III.*** Covariates

Covariates included socio-demographics, health status, and psychological factors that could influence difficulties during doctor visits. Socio-demographics encompassed gender, age (65-74, 75+), education (below high school, high school/trade school, some college or above), race/ethnicity (Black non-Hispanic, white non-Hispanic, Hispanic, other), and marital status (married/partnered or other). Health status covered medication management, vision impairment, hearing impairment, overall health, and having a regular doctor. Health conditions (e.g., heart disease, diabetes, arthritis) were scored from 0-10 and categorized as mild (1-2) or major (3-10). Impairments in activities of daily living (ADL) and instrumental activities of daily living (IADL) were also assessed. Psychological factors included self-rated memory (good to poor) and outlook on life, which was scored from 1-21 based on responses to seven questions. Scores were divided into mild (1-14) and highly positive outlook (15-21). Correlation analysis showed no significant relationships between the variables. The covariates for the presence or absence of hearing and vision loss were defined as follows. Hearing loss was assessed by asking participants if they had ever used a hearing aid or, for non-deaf individuals, whether they could perform certain tasks.

Hearing loss was recorded if they answered "no" to any of the following: (i) using a telephone, (ii) carrying on a conversation with a TV or radio playing, or (iii) holding a conversation in a quiet room. Vision loss was categorized into near and distance vision loss. Near vision loss was identified if participants could not read newspaper print or were blind, while distance vision loss was noted if they reported blindness or were unable to see across the street or view the TV across a room with glasses.

Table 1 below outlines all the covariates. For categorical variables, it includes their levels, frequencies, and percentages for Rounds 5 and 11. For numerical variables, it presents the means and standard deviations for both rounds. For the small portion of missing data, categorical variables were imputed using the mode, and numerical variables were imputed using the median.

The composite outcome variable indicating issues in the doctor’s office was analyzed using regression with all covariates. Initially, a bivariate mixed-effects binary logistic model was applied, regressing the outcome on each covariate individually. This was followed by a multivariate (adjusted) mixed-effects binary logistic regression that included all covariates at once. Weights were then incorporated into the adjusted model, and the analysis was redone.

Following this, the same analytical process was performed for each of the four questions that made up the outcome variable.

## Results

### ***I.*** Bivariate Analyses of Composite Outcome Variable

In the bivariate analysis, each covariate was assessed against the outcome variable using a mixed-effects binary logistic model, as shown in Table 2. Most covariates were significant predictors, except for "Other Race" compared to "White, non-Hispanic" (p-value = 0.139). Gender (p = 0.026) and marital status (p = 0.0161) reached significance at the 5% level, while other covariates were significant at the 0.001 level.

**Table 2.**
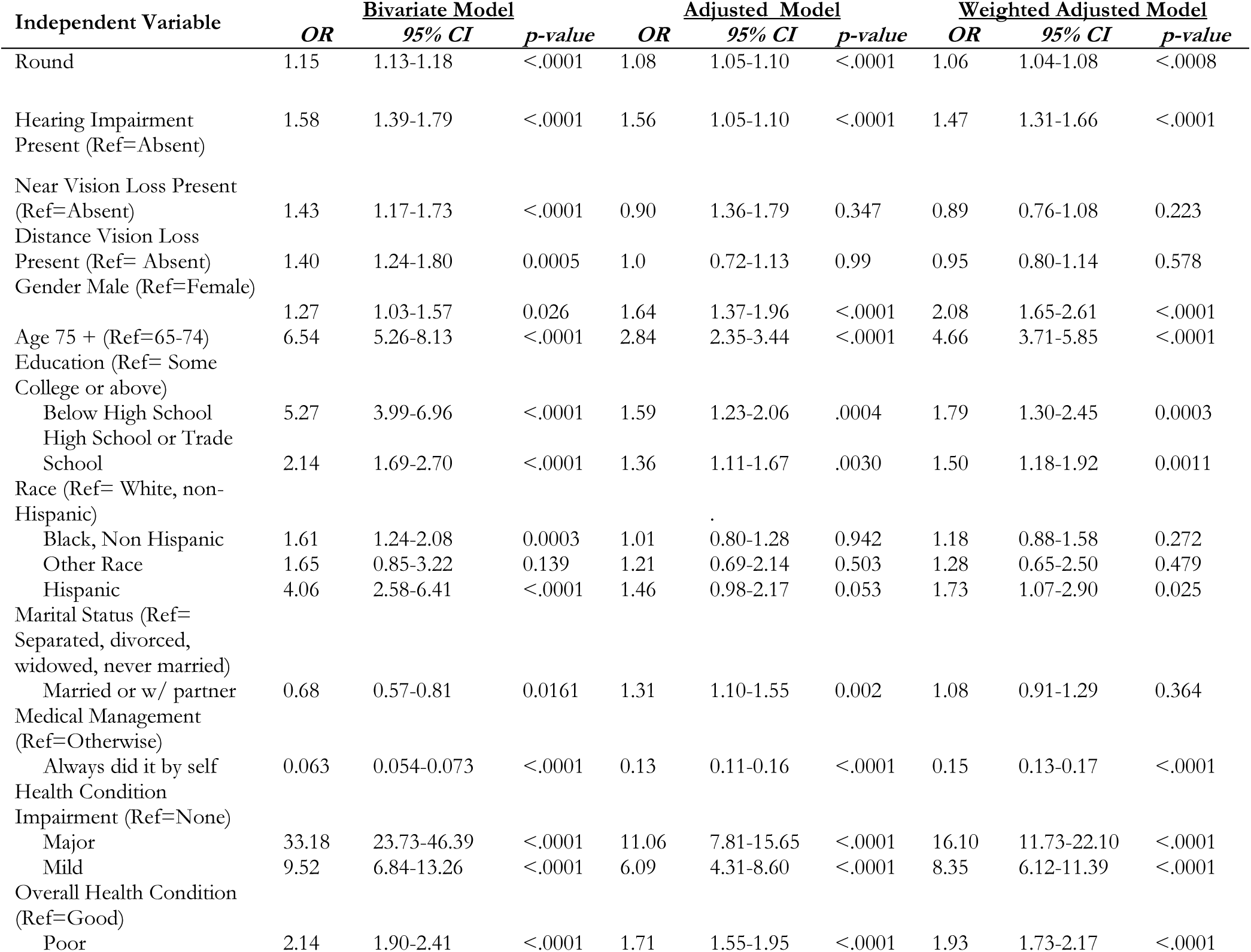
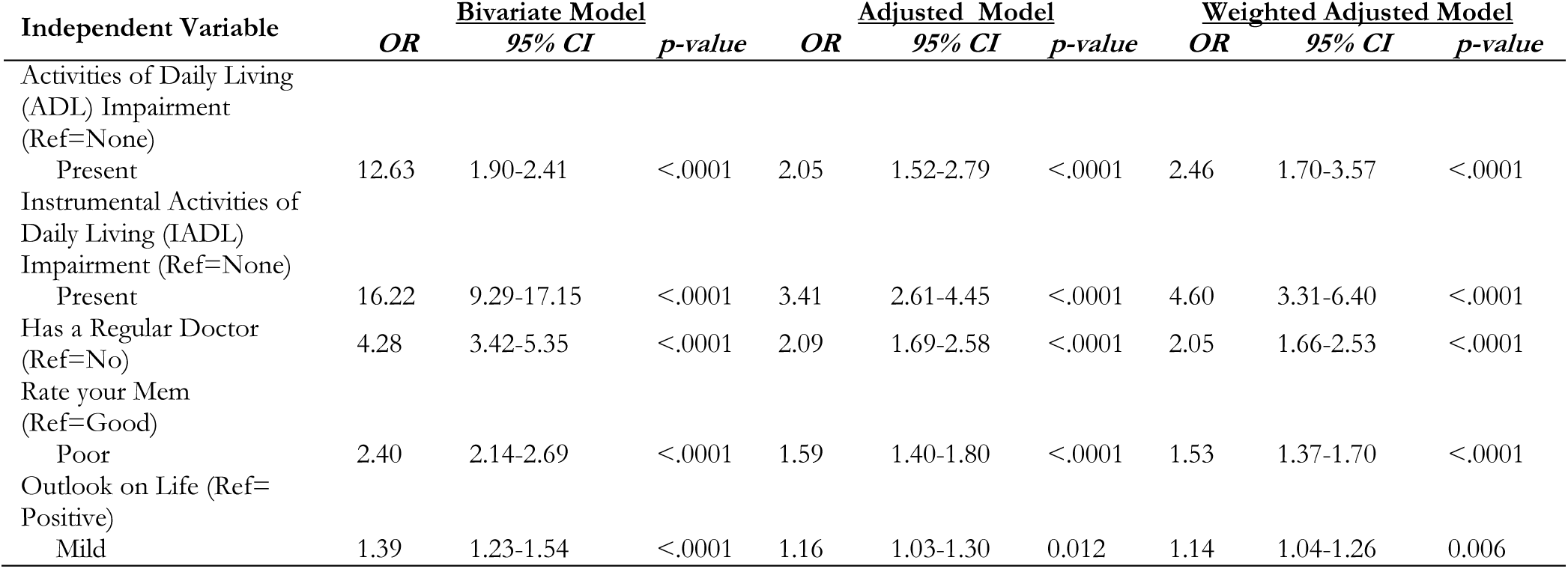
Results of bivariate and multivariate mixed-effects binary logistic regression analysis of composite outcome variable.

The highest odds ratios were observed for severe health condition impairments vs. none (OR: 33.18, 95% CI: 23.73–46.39), IADL impairments (OR: 16.22, 95% CI: 9.29–17.15), and ADL impairments (OR: 12.63, 95% CI: 9.29–17.15). Additional predictors included mild health condition impairments (OR: 9.52, 95% CI: 6.84–13.26), hearing impairment (OR: 1.58, 95% CI: 1.39–1.79), near vision loss (OR: 1.43, 95% CI: 1.17–1.73), and distance vision loss (OR: 1.40, 95% CI: 1.24–1.80). Educational level also showed a strong effect, with below high school education (OR: 5.27, 95% CI: 3.99–6.96) and high school or trade school education (OR: 2.14, 95% CI: 1.69–2.70) significantly increasing the odds of the outcome. Race effects included non-Hispanic Black (OR: 1.61, 95% CI: 1.24–2.08) and Hispanic (OR: 4.06, 95% CI: 2.58–6.41).

Conversely, being married or with a partner was protective (OR: 0.68, 95% CI: 0.57–0.81).

### ***II.*** Multivariate Analyses of Composite Outcome Variable

In the multivariate (adjusted) analysis, the mixed-effects binary logistic regression model revealed that most covariates retained significance, though some diminished in strength relative to the bivariate results. The detailed results in Table 2 show that near vision loss (p = 0.347), distance vision loss (p = 0.99), and all racial categories did not reach significance. Significant predictors with adjusted odds ratios include hearing impairment (OR = 1.56, 95% CI: 1.05– 1.10), education below high school (OR = 1.59, 95% CI: 1.23–2.06), and education at the high school or trade school level (OR = 1.36, 95% CI: 1.11–1.67). Marital status (married/partnered) showed a slightly elevated risk (OR = 1.31, 95% CI: 1.10–1.55), which contrasts with its protective effect in the bivariate analysis.

For health-related predictors, the odds ratio for major health condition impairments decreased but remained substantial (OR = 11.06, 95% CI: 7.81–15.65), with mild impairments also maintaining significance (OR = 6.09, 95% CI: 4.31–8.60). Functional impairments continued to play a significant role, with ADL impairment (OR = 2.05, 95% CI: 1.52–2.79) and IADL impairment (OR = 3.41, 95% CI: 2.61–4.45) contributing notably to the outcome.

The weighted adjusted regression analysis further clarifies the relationships between key covariates and the outcome variable. This analysis, using a mixed-effects binary logistic model, highlighted consistent non-significance for near vision loss (p = 0.223), distance vision loss (p = 0.578), and racial categories Black (p = 0.272) and Other Race (p = 0.479). Notably, Hispanic race became significant in the weighted model (OR: 1.73, 95% CI: 1.07-2.90), while marital status, previously significant in the unweighted model, was not (p = 0.364).

Significant predictors and their odds ratios in the weighted analysis include hearing impairment (OR = 1.47, 95% CI: 1.31–1.66), education below high school (OR = 1.79, 95% CI: 1.30–2.45), high school or trade school education (OR = 1.36, 95% CI: 1.11–1.67), and marital status, which now posed a slightly elevated risk (OR = 1.50, 95% CI: 1.18–1.92). Health condition impairments retained high odds ratios, with major impairments (OR = 16.10, 95% CI: 11.73– 22.10) and mild impairments (OR = 8.35, 95% CI: 6.12–11.39) as significant predictors.

Functional impairments also showed strong effects, with ADL impairment (OR = 2.46, 95% CI: 1.70–3.57) and IADL impairment (OR = 4.60, 95% CI: 3.31–6.40) contributing notably.

### ***III.*** Bivariate and Multivariate Analyses of Individual Questions

In this section we present the analyses conducted for each individual question.

#### 1. "In the last year, did anyone sit in with (you/SP) and (your/(their))(regular) doctor during (your/his/her) visits?

In the bivariate analysis, each covariate was assessed against the outcome variable using a mixed-effects binary logistic model, as shown in Table 3. Most covariates were significant predictors, except for "Distance Vision Loss" (p-value=0.09) and "Black, non-Hispanic" (p-value = 0.26), yielding the highest p-value of the covariates in the group "Other Race" (p=0.52). Gender (p = 0.03) reached significance at the 5% level, while other covariates were significant at the 0.001 level.

**Table 3.** Results of mixed-effects binary logistic regression analyses for question “ *In the last year, did anyone sit in with (you/SP) and (your/(their))(regular) doctor during (your/his/her) visits?*”

| Independent Variable | Bivariate Model |  |  | Adjusted Model |  |  | Weighted Adjusted Model |  |  |
| --- | --- | --- | --- | --- | --- | --- | --- | --- | --- |
|  | OR | 95% CI | p-value | OR | 95% CI | p-value | OR | 95% CI | p-value |
| Round | 1.08 | 1.06-1.10 | <.0001 | 1.05 | 1.03-1.07 | <.0001 | 1.02 | 1.01-1.04 | 0.01 |
| Hearing Impairment Present (Ref=Absent) | 1.22 | 1.09-1.36 | <.0001 | 1.31 | 1.16-1.48 | <.0001 | 1.26 | 1.14-1.40 | <.0001 |
| Near Vision Loss Present (Ref=Absent) | 1.17 | 0.97-1.41 | 0.0007 | 0.93 | 0.75-1.14 | 0.48 | 0.96 | 0.80-1.15 | 0.631 |
| Distance Vision Loss Present (Ref= Absent) | 1.35 | 1.14-1.61 | 0.09 | 1.18 | 0.97-1.43 | 0.091 | 1.17 | 0.99-1.38 | 0.067 |
| Gender Male (Ref=Female) | 1.22 | 1.02-1.46 | 0.03 | 1.21 | 1.02-1.43 | 0.03 | 1.36 | 1.11-1.66 | 0.003 |
| Age 75 + (Ref=65-74) | 4.83 | 3.79-6.16 | <.0001 | 3.02 | 2.55-3.57 | <.0001 | 4.14 | 3.40-5.03 | <.0001 |
| Education (Ref= Some College or above) |  |  |  |  |  |  |  |  |  |
| Below High School | 3.10 | 2.46-3.93 | <.0001 | 1.61 | 1.28-2.04 | <.0001 | 1.75 | 1.32-2.31 | <.0001 |
| High School or Trade School | 1.62 | 1.33-1.97 | <.0001 | 1.24 | 1.04-1.48 | 0.020 | 1.35 | 1.09-1.67 | 0.006 |
| Race (Ref= White, non-Hispanic) |  |  |  |  |  |  |  |  |  |
| Black, Non Hispanic | 1.14 | 0.91-1.41 | 0.26 | 0.93 | 0.75-1.15 | 0.48 | 0.98 | 0.76-1.27 | 0.865 |
|  | OR | 95% CI | p-value | OR | 95% CI | p-value | OR | 95% CI | p-value |
| Other Race | 1.21 | 0.98-2.15 | 0.52 | 1.03 | 0.61-1.73 | 0.906 | 1.12 | 0.61-2.03 | 0.723 |
| Hispanic | 2.43 | 1.64-3.62 | <.0001 | 1.26 | 0.87-1.83 | 0.229 | 1.33 | 0.86-2.04 | 0.201 |
| Marital Status (Ref= Separated, divorced, widowed, never married) |  |  |  |  |  |  |  |  |  |
| Married or w/ partner | 1.56 | 1.35-1.79 | <.0001 | 2.77 | 2.40-3.21 | <.0001 | 2.77 | 2.40-3.18 | <.0001 |
| Medical Management (Ref=Otherwise) |  |  |  |  |  |  |  |  |  |
| Always did it by self | 0.09 | 0.08-0.11 | <.0001 | 0.17 | 0.14-0.20 | <.0001 | 0.20 | 0.18-0.23 | <.0001 |
| Health Condition Impairment (Ref=None) |  |  |  |  |  |  |  |  |  |
| Major | 29.05 | 22.48-37.53 | <.0001 | 13.45 | 10.29-17.57 | <.0001 | 14.61 | 11.68-18.28 | <.0001 |
| Mild | 9.34 | 7.27-11.99 | <.0001 | 6.44 | 4.96-8.36 | <.0001 | 5.99 | 4.84-7.40 | <.0001 |
| Overall Health Condition (Ref=Good) |  |  |  |  |  |  |  |  |  |
| Poor | 1.57 | 1.41-1.75 | <.0001 | 1.50 | 1.33-1.69 | <.0001 | 1.56 | 1.41-1.72 | <.0001 |
| Activities of Daily Living (ADL) Impairment (Ref=None) |  |  |  |  |  |  |  |  |  |
| Present | 7.49 | 5.71-9.83 | <.0001 | 1.95 | 1.47-2.58 | <.0001 | 2.15 | 1.53-3.01 | <.0001 |
| Instrumental Activities of Daily Living (IADL) Impairment (Ref=None) |  |  |  |  |  |  |  |  |  |
| Present | 8.77 | 6.94-10.09 | <.0001 | 2.70 | 2.10-3.48 | <.0001 | 3.46 | 2.56-4.66 | <.0001 |
| Has a Regular Doctor (Ref=No) | 4.21 | 3.52-5.05 | <.0001 | 2.07 | 1.69-2.55 | <.0001 | 1.68 | 1.43-1.98 | <.0001 |
| Rate your Memory (Ref=Good) |  |  |  |  |  |  |  |  |  |
| Poor | 1.38 | 1.25-1.53 | <.0001 | 1.06 | 0.95-1.19 | 0.301 | 1.02 | 0.93-1.12 | 0.70 |
| Outlook on Life (Ref= Positive) |  |  |  |  |  |  |  |  |  |
| Mild | 1.35 | 1.23-1.49 | <.0001 | 1.14 | 1.04-1.26 | 0.008 | 1.07 | 0.99-1.16 | 0.09 |

*2. "Did you or anyone remind SP of questions you wanted to ask the doctor during the doctor's visit last year?"*

The highest odds ratios were observed for severe health condition impairments vs. none (OR: 29.06, 95% CI: 22.48-37.53), mild health condition impairments vs none (OR: 9.34, 95% CI: 7.27-11.99)IADL impairments (OR: 8.77, 95% CI: 6.94-10.09), and ADL impairments (OR: 7.49, 95% CI:5.71-9.83). In terms of sensory factors, hearing impairment (OR: 1.22, 95% CI: 1.09-1.36), near vision loss (OR: 1.17, 95% CI: 0.97-1.41), and distance vision loss (OR: 1.35, 95% CI: 1.14-1.61). Education level had some, but not too high of an effect, with below high school education (OR: 3.10, 95% CI: 2.46-3.93) and high school or trade school education (OR: 1.62, 95% CI: 1.33-1.97) showing moderate, to little, increases in odds ratios. Age (OR: 4.83, 95% CI: 3.79-6.16) significantly impacted participants’ responses to this question. Older participants may need assistance during doctor visits. Race effects included non-Hispanic Black (OR: 1.14, 95% CI: 0.91-1.41) and Hispanic (OR: 1.21, 95% CI:1.64-3.62). Conversely, being married or with a partner was protective (OR: 1.58, 95% CI: 1.35-1.79).

In the multivariate-adjusted analysis, the mixed-effects binary logistic model revealed decreases in odds ratios for most, if not all, of the covariates; however, a few increased in odds ratios during the transition. The results in Table 3 show that near vision loss increased in insignificance (p=0.48) as well as in all racial categories. Significant covariates in this multivariate model include hearing impairment (OR=1.31, 95% CI: 1.16-1.48), Education below high school (OR=1.61, 95% CI: 1.28-2.04), education at the high school or trade level (OR= 1.24, 95% CI: 1.04-1.48). Marital status (OR=2.77, 95% CI: 2.40-3.21) contributed significantly to this question, with its odds ratio increasing considerably more than in the bivariate analysis.

Concerning the medical and sensory factors, we noted the odds ratio for significant health condition impairment decreased by more than 50% (OR: 13.4, 95% CI: 10.29-17.57) while minor health condition imprisonment decreased slightly (OR: 6.44, 95% CI: 4.96-8.36) as we transition from bivariate to multivariate. Similarly, ADL and IADL impairments showed significant drops in odds ratios (1.95 and 2.70, respectively), yet still positive and significant, contributing to the questionnaire response.

As we ran our weighted adjusted regression model, we discovered that Near vision loss (p=0.631) and distance vision loss (p=0.067), as well as all racial categories, remained insignificant. Compared to the adjusted model, all covariates of the weighted model still retained their significance or insignificance, and no one variable changed during this transition.

The significant predictors and their odds ratios in this weighted model were hearing impairment (OR: 1.26, 95% CI: 1.14-1.40), Education below high school (OR: 1.75, 95% CI: 1.32-2.31), education at either the high school or trade level (OR: 1.35, 95% CI: 1.09-1.67) and with the same odds ratio as the multivariate model, marital status (OR: 2.77, 95% CI: 2.40-3.18) which still contributes significantly to influencing question response. Health condition impairments still yielded the highest odds ratios, with significant (OR: 14.61, 95% CI: 11.68-18.28) and mild (5.99, 95% CI: 4.84-7.40) yielding the highest values in the model. Both of these values are significant. Lastly, functional impairments were still deemed significant and contributing significantly to the response, with ADL (OR: 2.15, 95% CI: 1.53-3.01) and IADL (OR: 3.46, 95% CI: 2.56-4.66).

#### 2. “Did you or anyone remind SP of questions you wanted to ask the doctor during the doctor’s visit last year?”

In the bivariate analysis, each covariate was assessed against the outcome variable using a mixed-effects binary logistic model, as shown in Table 4. Most covariates were significant predictors, except for "Education below High School" (p-value=0.10) " Education at either the trade or high school" (p-value = 0.99) and "Marital Status" (p=0.47). Distance Vision Loss(p = 0.006) was significant at the 5% level, while other covariates were significant at the 0.001 level. The highest odds ratios were observed for severe health condition impairments vs none (OR: 23.55, 95% CI: 17.05-32.52), mild health condition impairments vs none (OR: 7.31, 95% CI: 5.31-10.07) ADL impairments (OR: 8.23, 95% CI: 6.12-11.07), and IADL impairments (OR: 10.98, 95% CI:8.49-14.21). In terms of sensory factors, hearing impairment (OR: 1.46, 95% CI: 1.28-1.64), near vision loss (OR: 1.33, 95% CI: 1.09-1.62), and distance vision loss (OR: 1.53, 95% CI: 1.28-1.96). Education level still significantly contributed to response, with below high school education (OR: 3.11, 95% CI: 2.37-4.07) and high school or trade school education (OR: 1.63, 95% CI: 1.30-2.04). Race showed little to no association affecting questionnaire response, with non-Hispanic Black (OR: 1.23, 95% CI: 0.96-1.59) and Hispanic (OR: 2.64, 95% CI:1.69- 4.10). Likewise, being married or with a partner showed minimal association (OR: 1.06, 95% CI: 0.90-1.25).

**Table 4.** Results of mixed-effects binary logistic regression analyses for question “*Did you or anyone remind SP of questions you wanted to ask the doctor during the doctor’s visit last year?*

| Independent Variable | <u>Bivariate Model</u> |  |  | <u>Adjusted Model</u> |  |  | <u>Weighted Adjusted Model</u> |  |  |
| --- | --- | --- | --- | --- | --- | --- | --- | --- | --- |
|  | OR | 95% CI | p-value | OR | 95% CI | p-value | OR | 95% CI | p-value |
| Round | 1.12 | 1.09-1.14 | <.0001 | 1.06 | 1.03-1.08 | <.0001 | 1.05 | 1.03-1.07 | <.0001 |
| Hearing Impairment Present (Ref=Absent) | 1.46 | 1.28-1.64 | <.0001 | 1.39 | 1.22-1.59 | <.0001 | 1.41 | 1.26-1.59 | <.0001 |
| Near Vision Loss Present (Ref=Absent) | 1.33 | 1.09-1.62 | <.0001 | 0.86 | 0.96-1.08 | 0.20 | 0.82 | 0.67-0.99 | 0.043 |
| Distance Vision Loss Present (Ref= Absent) | 1.53 | 1.28-1.96 | 0.006 | 1.12 | 0.92-1.37 | 0.275 | 1.17 | 0.98-1.40 | 0.083 |
| Gender Male (Ref=Female) | 1.60 | 1.31-1.96 | <.0001 | 1.80 | 1.49-2.19 | <.0001 | 2.32 | 1.85-2.92 | <.0001 |
| Age 75 + (Ref=65-74) | 6.10 | 4.52-8.24 | <.0001 | 2.59 | 2.15-3.12 | <.0001 | 3.97 | 3.17-4.97 | <.0001 |
| Education (Ref= Some College or above) |  |  |  |  |  |  |  |  |  |

| Independent Variable | Bivariate Model |  |  | Adjusted Model |  |  | Weighted Adjusted Model |  |  |
| --- | --- | --- | --- | --- | --- | --- | --- | --- | --- |
|  | OR | 95% CI | p-value | OR | 95% CI | p-value | OR | 95% CI | p-value |
| Below High School | 3.11 | 2.37-4.07 | <.0001 | 1.22 | 0.95-1.59 | <.0001 | 1.31 | 0.96-1.80 | 0.094 |
| High School or Trade School | 1.63 | 1.30-2.04 | <.0001 | 1.17 | 0.96-1.43 | 0.073 | 1.26 | 0.99-1.61 | 0.061 |
| Race (Ref= White, non-Hispanic) |  |  |  |  |  |  |  |  |  |
| Black, Non Hispanic | 1.23 | 0.96-1.59 | 0.10 | 0.93 | 0.74-1.18 | 0.564 | 1.05 | 0.79-1.41 | 0.725 |
| Other Race | 1.00 | 0.52-1.93 | 0.99 | 0.73 | 0.41-1.30 | 0.289 | 0.76 | 0.39-1.51 | 0.434 |
| Hispanic | 2.64 | 1.69-4.10 | <.0001 | 1.18 | 0.79-1.79 | 0.427 | 1.36 | 0.84-2.19 | 0.215 |
| Marital Status (Ref= Separated, divorced, widowed, never married) |  |  |  |  |  |  |  |  |  |
| Married or w/ partner | 1.06 | 0.90-1.25 | 0.47 | 1.75 | 1.48-2.06 | <.0001 | 1.49 | 1.26-1.76 | <.0001 |
| Medical Management (Ref=Otherwise) |  |  |  |  |  |  |  |  |  |
| Always did it by self | 0.10 | 0.08-0.11 | <.0001 | 0.18 | 0.16-0.22 | <.0001 | 0.21 | 0.19-0.25 | <.0001 |
| Health Condition Impairment (Ref=None) |  |  |  |  |  |  |  |  |  |
| Major | 23.55 | 17.05-32.52 | <.0001 | 9.57 | 6.85-13.37 | <.0001 | 13.89 | 10.25-18.83 | <.0001 |
| Mild | 7.31 | 5.31-10.07 | <.0001 | 4.86 | 3.49-6.77 | <.0001 | 6.58 | 4.89-8.67 | <.0001 |
| Overall Health Condition (Ref=Good) |  |  |  |  |  |  |  |  |  |
| Poor | 1.93 | 1.71-2.17 | <.0001 | 1.62 | 1.42-1.84 | <.0001 | 1.74 | 1.56-1.94 | <.0001 |
| Activities of Daily Living (ADL) Impairment (Ref=None) |  |  |  |  |  |  |  |  |  |
| Present | 8.23 | 6.12-11.07 | <.0001 | 1.81 | 1.34-2.44 | <.0001 | 2.13 | 1.47-3.09 | <.0001 |
| Instrumental Activities of Daily Living (IADL) Impairment (Ref=None) |  |  |  |  |  |  |  |  |  |
| Present | 10.98 | 8.49-14.21 | <.0001 | 3.10 | 2.37-5.05 | <.0001 | 4.17 | 3.00-5.81 | <.0001 |
| Has a Regular Doctor (Ref=No) | 3.92 | 3.14-4.89 | <.0001 | 2.38 | 1.85-3.06 | <.0001 | 1.87 | 1.52-2.29 | <.0001 |
| Rate your Memory (Ref=Good) |  |  |  |  |  |  |  |  |  |
| Poor | 1.96 | 1.75-2.20 | <.0001 | 1.38 | 1.22-1.56 | <.0001 | 1.35 | 1.21-1.50 | <.0001 |
| Outlook on Life (Ref= Positive) |  |  |  |  |  |  |  |  |  |
| Mild | 1.41 | 1.27-1.57 | <.0001 | 1.22 | 1.10-1.37 | 0.0004 | 1.22 | 1.11-1.34 | <.0001 |

In the multivariate-adjusted analysis, the mixed-effects binary logistic model revealed decreases in odds ratios for most, if not all, of the covariates; however, a few increased in odds ratios during the transition. The results in Table 4show that near-vision loss increased in insignificance (p=0.20). Distance vision loss (p=0.275) and all racial categories are the same. Significant covariates in this multivariate model include hearing impairment (OR=1.39, 95% CI: 1.22-1.59), Education below high school (OR=1.22, 95% CI: 0.95-1.59), education at the high school or trade level (OR= 1.17, 95% CI: 0.96-1.43). Marital status (OR=1.75, 95% CI: 1.48-2.06) contributed moderately to this question, with its odds ratio just slightly larger than that of the bivariate model.

Concerning the medical and sensory factors, the odds ratio for major health condition impairment decreased significantly (OR: 9.57, 95% CI: 6.85-13.37), while minor health condition imprisonment decreased moderately (OR: 4.86, 95% CI: 3.49-6.77) from bivariate to multivariate. The p-values for the preceding covariates remain significant. Likewise, ADL and IADL impairments showed major drops in odds ratios (1.81 and 3.10, respectively), yet still positive and vital, contributing to the questionnaire response. However, ADL will likely have a negligible influence compared to IADL.

As we ran our weighted adjusted regression model, we discovered that at the 5% significance level, Near vision loss (p=0.043) was significant, but distance vision loss (p=0.083) wasn’t. This is also true for racial categories, as they remain insignificant. Apart from near and distance vision loss, all covariates of the weighted model still retained their original significance.

The significant predictors and their odds ratios in this weighted model were hearing impairment (OR: 1.41, 95% CI: 1.26-1.59), Education below high school (OR: 1.31, 95% CI: 0.96-1.80), education at either the high school or trade level (OR: 1.26, 95% CI: 0.99-1.61). Marital status (OR: 1.49, 95% CI: 1.26-1.76) significantly increased odds ratios during the transition. Health condition impairments still yielded the highest odds ratios with central (OR: 13.89, 95% CI: 10.25-18.83) and mild (6.58, 95% CI: 4.89-8.67) yielding the highest values within the model, both of these values are significant. Lastly, functional impairments were still deemed significant and contributing significantly to the response, with ADL (OR: 2.13, 95% CI: 1.47-3.09) and IADL (OR: 4.17, 95% CI: 3.00-5.81).

#### 3. “During doctor visits last year, did you or anyone ask/tell things for you/sp?”

In the bivariate analysis, each covariate was assessed against the outcome variable using a mixed-effects binary logistic model, as shown in Table 5. In our review, all but two covariates came out significant. These two insignificant covariates are Male (p=0.15) and "Other Race" (p=0.11). The remaining covariates were significant at the 0.001 level.

**Table 5.**
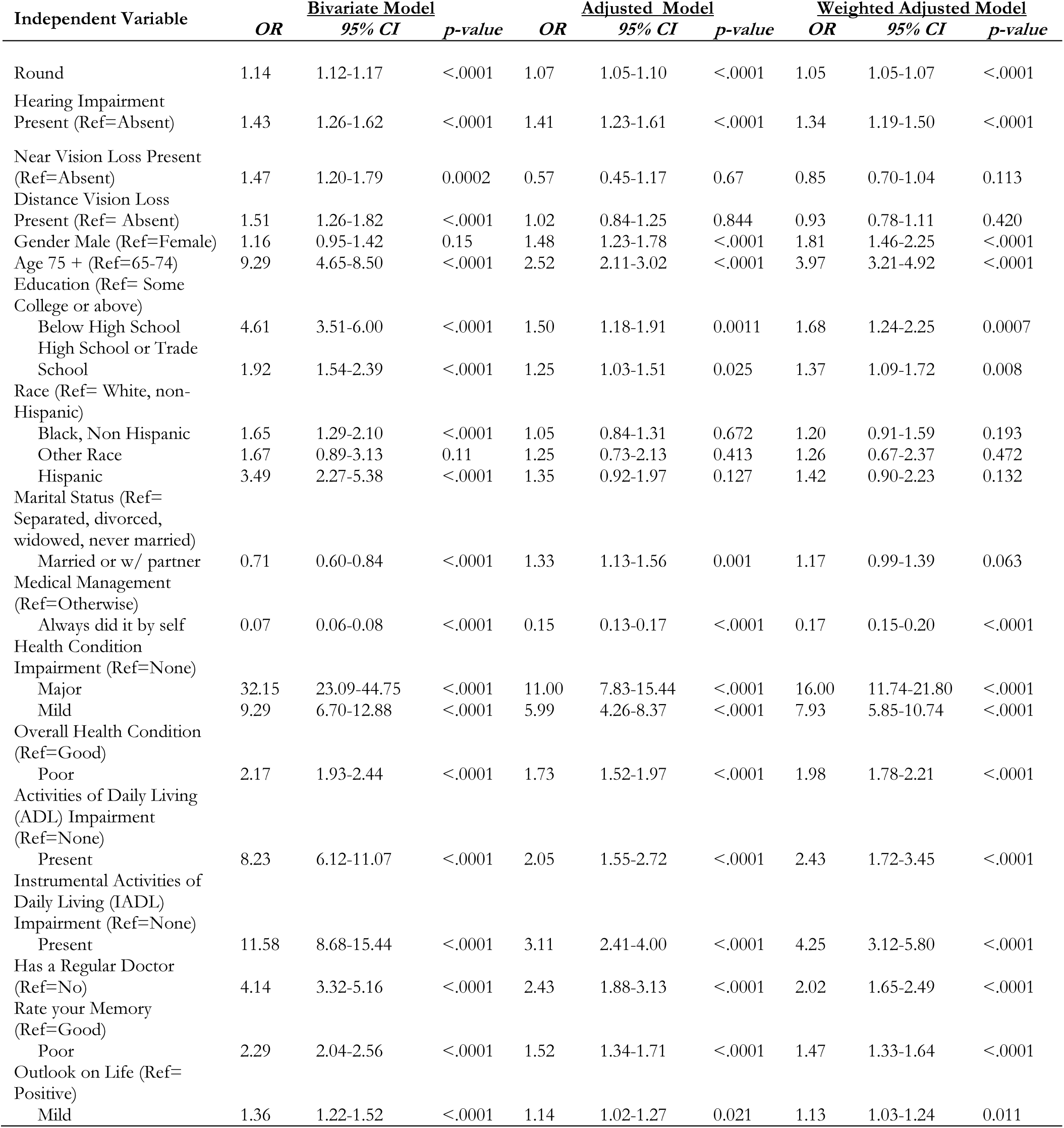
Results of mixed-effects binary logistic regression analyses for question “*During doctor visits in the last year, did you or anyone ask/tell things for you/SP?*”

The highest odds ratios were observed for severe health condition impairments vs. none (OR: 11.00, 95% CI: 7.83-15.4), mild health condition impairments vs none (OR: 5.99, 95% CI: 4.26-8.37), IADL impairments (OR: 3.11, 95% CI: 2.41-4.00), and ADL impairments (OR: 2.05, 95% CI:1.55-2.72). In terms of sensory factors, hearing impairment (OR: 1.41, 95% CI: 1.23-1.61) showed a small but positive association, distance vision loss (OR: 1.02, 95% CI: 0.84-1.25) displayed minuscule associations, and near vision loss (OR: 0.57, 95% CI: 0.45-1.17) indicated a declining association, suggesting vision plays no factor in influencing participant response for this particular question. Education level had minor effects, with below high school education (OR: 1.50, 95% CI: 1.18-1.91) and high school or trade school education (OR: 1.25, 95% CI: 1.03-1.51) showing, to minor, increases in association. Race effects included non-Hispanic Black (OR: 1.05, 95% CI: 0.84-1.31) and Hispanic (OR: 1.35, 95% CI:0.92-1.97), and Other race (OR: 1.25, 95% CI: 0.73-2.13), all racial covariates are insignificant. Conversely, being married or with a partner was protective (OR: 1.33, 95% CI: 1.13-1.56).

In the multivariate-adjusted analysis, the mixed-effects binary logistic model revealed decreases in odds ratios for most, if not all, of the covariates; however, a few increased in odds ratios during the transition. The results in Table 5 show that near vision loss (p=0.48) and distance vision loss (p=0.84) increased in insignificance as well as in all racial categories. Education at the high school or trade school (p=0.025) increased marginally in insignificance but not enough to reach the 5% level. This is also true for Education below high school (OR=1.50, 95% CI: 1.18-1.91) where p=0.0011. Significant covariates in this multivariate model include hearing impairment (OR=1.41, 95% CI: 1.23-1.61). Marital status (OR=1.33, 95% CI: 1.13-1.56) contributed significantly to this question, with its odds ratio increasing considerably more than in the bivariate analysis.

Concerning the medical and sensory factors, we noted the odds ratio for significant health condition impairment decreased by more than 66% (OR: 11.00, 95% CI: 7.83-15.44) while minor health condition imprisonment decreased slightly (OR: 5.99, 95% CI: 4.26-8.37) as we transition from bivariate to multivariate. Similarly, ADL and IADL impairments showed significant drops in odds ratios (2.05 and 3.11, respectively) yet remained high, positively significantly contributing to the questionnaire response.

As we ran our weighted adjusted regression model, we noted that Near vision loss (p=0.113) and distance vision loss (p=0.420) were still insignificant; however, the odds ratios were <1, indicating a negative association as vision loss did not influence this hearing question. Aside from that, all racial categories remained insignificant. Compared to the adjusted model, all covariates of the weighted model still retained their significance, and many of them increased in odds ratios ever so slightly.

At the 5% level, The significant predictors and their odds ratios in this weighted model were hearing impairment (OR: 1.34, 95% CI: 1.19-1.50), Education below high school (OR: 1.68, 95% CI: 1.24-2.25), education at either the high school or trade level (OR: 1.37, 95% CI:1.09- 1.72). Crossing above the 5% level, marital status (OR: 1.17, 95% CI: 0.99-1.39) became insignificant but still yielded a positive association. Health condition impairments still yielded the highest odds ratios, with significant (OR: 16.00, 95% CI: 11.74-21.80) and mild (7.93, 95% CI: 5.85-10.74) yielding the highest values in the model. Both of these values are significant.

Lastly, functional impairments were still deemed significant and contributing significantly to the response, with ADL (OR: 2.43, 95% CI: 1.72-3.45) and IADL (OR: 4.25, 95% CI: 3.12-5.80).

#### 4. “During those visits in the last year, did you or anyone else help SP understand what the doctor was saying?”

In the bivariate analysis, each covariate was assessed against the outcome variable using a mixed-effects binary logistic model, as shown in Table 6. At the 0.001 level, most covariates were significant predictors, except for "Near Vision Loss" (p-value=00029) "Other Race" (p-value = 0.12) and yielding the highest p-value of the covariates in the group "Other Race" (p=0.52). Gender (p = 0.54) was insignificant, and other covariates were nonetheless significant. The highest odds ratios were observed for severe health condition impairments vs. none (OR: 32.74, 95% CI: 22.63-44.52), mild health condition impairments vs none (OR: 9.14, 95% CI: 6.55-12.76), IADL impairments (OR: 17.37, 95% CI: 13.28-22.72), and ADL impairments (OR: 14.10, 95% CI:10.32-19.26). In terms of sensory factors, hearing impairment (OR: 1.68, 95% CI: 1.47-1.91), near vision loss (OR: 1.36, 95% CI: 1.11-1.67), and distance vision loss (OR: 1.57, 95% CI: 1.30-1.90). Education, to our surprise, has a significant effect on questionnaire responses where Below High School(OR: 7.91, 95% CI: 5.94-10.50) and high school or trade school education (OR: 2.67, 95% CI: 2.11-3.38) with its high odds ratios. This suggests that those with low education levels may encounter difficulty understanding basic English, contributing to problems with doctor visits. Race effects included non-Hispanic Black (OR: 1.94, 95% CI: 1.49-2.52) and Hispanic (OR: 5.57, 95% CI:3.51-8.83). The participants’ low education level and Hispanic heritage [19] may have contributed to difficulties understanding spoken English conversation with the doctor, necessitating communication aid. Marital status, on the other hand, yielded a declining association as indicated by its odds ratio (OR: 0.56, 95% CI: 0.47-0.67).

**Table 6.**
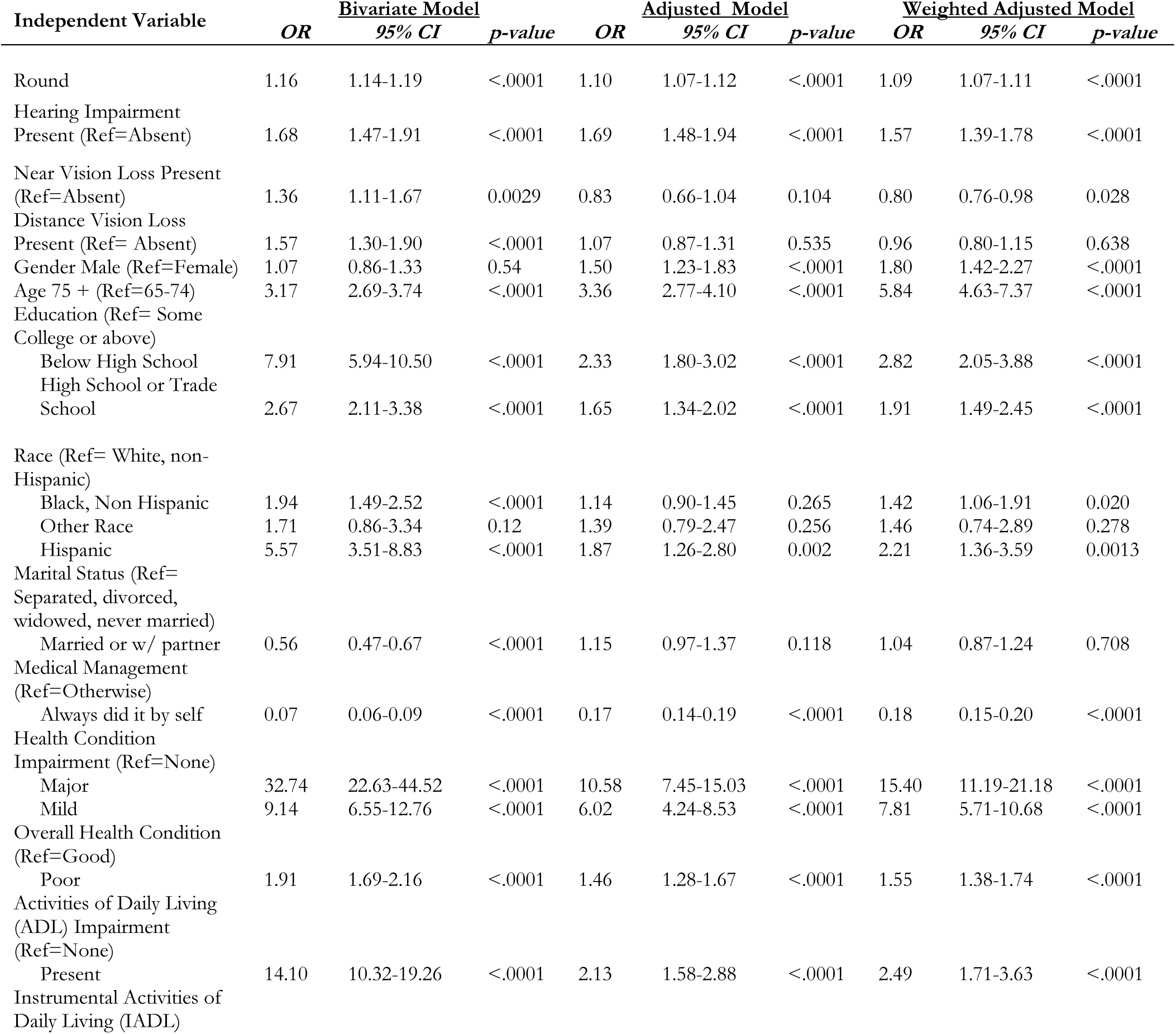
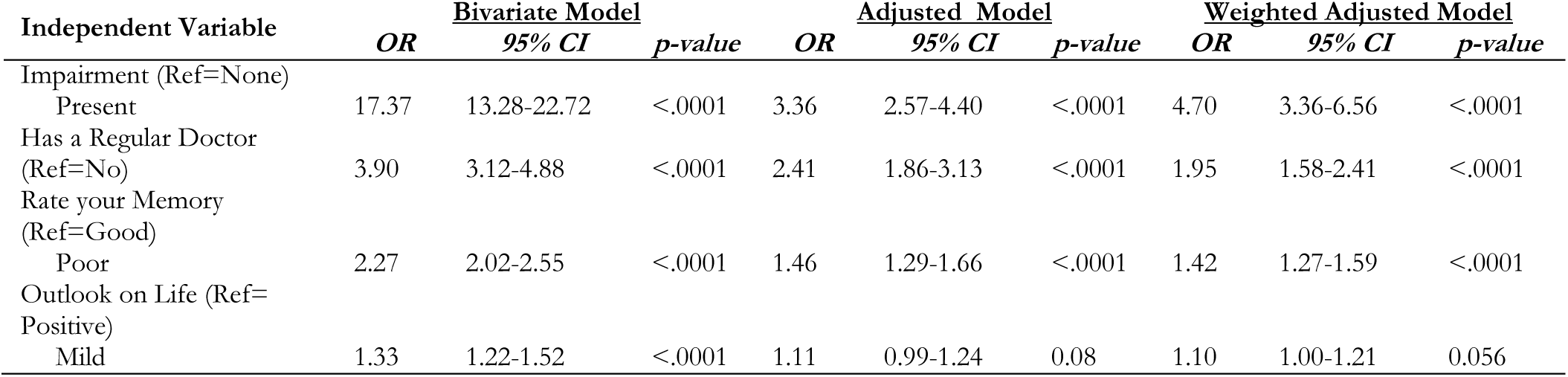
Results of mixed-effects binary logistic regression analyses for question “During those visits in the last year, did you or anyone else help SP understand what the doctor was saying?”

In the multivariate-adjusted analysis, the mixed-effects binary logistic model revealed decreases in odds ratios for most, if not all, covariates; however, a few increased in odds ratios during the transition. The results in Table 6 show that near vision loss(p=0.104) and distance vision loss (p=0.535) increased in insignificance as well as all racial categories (except HISPANIC, as that remained significant). Significant covariates in this multivariate model include hearing impairment (OR=1.69, 95% CI: 1.48-1.94), Education below high school (OR=2.33, 95% CI: 1.80-3.02), education at the high school or trade level (OR= 1.65, 95% CI: 1.34-2.02). Marital status (OR=1.15, 95% CI: 0.97-1.37), despite having a favorable odds ratio, became more insignificant (p=0.118), indicating that having a partner really did not really help the participant understand what the doctor was saying.

Concerning the medical and sensory factors, we noted the odds ratio for significant health condition impairment decreased by more than 60% (OR: 10.58, 95% CI: 7.45-15.03) while minor health condition imprisonment decreased slightly (OR: 6.02, 95% CI: 4.24-8.53) as we transition from bivariate to multivariate. Similarly, ADL and IADL impairments showed significant drops in odds ratios (2.13 and 3.36, respectively), yet still positive and significant, contributing to the questionnaire response.

As we ran our weighted adjusted regression model, Near vision loss (p=0.028) was insignificant at the 0.001 level yet still significant at the 5% level. Distance vision loss (p=0.638) retained its insignificance, and all racial categories (with Hispanic being the exception) remained significant during this transition. The significant predictors and their odds ratios in this weighted model were hearing impairment (OR: 1.57, 95% CI: 1.39-1.78), Education below high school (OR: 2.82, 95% CI: 2.05-3.88), education at either the high school or trade level (OR: 1.91, 95% CI: 1.49-2.45). Health condition impairments resulted in high odds ratios with central (OR: 15.40, 95% CI: 11.19-21.18) and mild (OR: 7.81, 95% CI: 5.71-10.68) yielding the highest values in the model. Both of these values are significant. Lastly, functional impairments were still deemed significant and contributing significantly to the response, with ADL (OR: 2.49, 95% CI: 1.71- 3.63) and IADL (OR: 4.70, 95% CI: 3.36-6.56)

## Discussion

This observational study analyzed data from the NHATS (Round 5 –11) to identify factors influencing doctor-patient communication in a nationally representative sample of community-dwelling older adults. Our findings revealed significant predictors at the demographic, sensory/functional, health/health care, medication management, perceived cognitive health, and self-efficacy levels. Regarding demographics, we found two main factors contributing to communication difficulties: being male or Hispanic. Males were twice as likely to have a companion present during a doctor’s visit compared to their female counterparts, highlighting the need for support with communication disabilities. This is supported by relevant literature. Adults above 50 years and women reported higher patient-centered provider communication. However, Hispanic and Asian respondents reported poorer patient-centered provider communication as compared to their White counterparts. [11, 12]. In addition to racial demographics, we observed that participants with a low educational level below high school significantly required more assistance. Accompanied older adults who received task assistance were often older, less educated, and had worse self-rated health [14].

Regarding sensory and functional limitations, participants who reported hearing impairment (refer to subsection III of the methods section) yielded significant communication difficulties, despite using a hearing aid. Regardless, hearing loss yielded minimal associations with improvements in hearing-related function [2]. This loss of sensory impairment can lead to higher associations of self-care adverse consequences, such as depression [7]. When comparing vision loss to hearing impairment, a higher percentage of Medicare beneficiaries with hearing impairment indicated communication-related reasons required assistance in oral communication. [8].

In addition, difficulties in task performance (ADL or IADL) due to a health or physical condition often require a companion present for assistance [14]. Interestingly, multiple chronic health conditions were identified as our model’s largest and most significant contributing factor. Our results indicate that having at least one major chronic condition significantly impacted the patients’ difficulty at the medical office. Recent literature argued that the elderly with both chronic health conditions had ineffective communication with their doctor as compared to those with only acute illnesses [3]. Highlighting the more detrimental and long-lasting effects of chronic conditions on social relationships compared to acute diseases.

While most of the factors discussed in this article focus on the negative aspects affecting communication, we have noted several positives within our study. Psychologically, participants who have reported confidence through a positive outlook on life and excellent health predicted fewer issues with the doctor [12]. Those continuously seeking help view their healthcare providers as helpful, while non-help-seekers may think they will not be taken seriously. Our results illustrate the importance of the patient-provider relationship in creating a welcoming atmosphere where older adults are comfortable and confident, promoting positive cognitive function and well-being [1].

Our study has several limitations. First, we are working with a limited dataset not typically representative of the overall NHATS population. Due to the lack of responses from many participants in rounds 5-11, we excluded them from the dataset, reducing the original sample size from approximately 12,000 to 3,700 participants. Some of these excluded participants may have provided important information through their responses that could have enhanced our analysis.

Second, among the participants who did respond to all questions in rounds 5-11, some provided missing or inapplicable responses to specific questionnaire questions that were crucial for our research. These missing responses were due to factors beyond our control. To address this, we had to treat these responses as "no" or use median imputation for non-numeric categorical responses. This approach may introduce bias and uncertainty into the data, potentially affecting the results of our model. Future research should address missing or incomplete data to ensure high-quality, more consistent, and diverse results. Third, we have a limited number of variables in our dataset. While the selected variables align with our research objectives, we could have included additional variables to strengthen our model. Incorporating health behavior-related predictors such as smoking, physical exercise intensity, sleeping habits, and weight loss progress could provide valuable insights. However, adding these variables might not align with our hypothesis and could lead to redundant observations. Further research is needed to explore the impact of these additional variables on our model’s performance.

Despite our limitations, we have identified several key strengths within our study. First, we analyzed our data using multiple statistical techniques, including regression analysis. This approach allowed us to explore the complex relationships between variables, leading to evidence for our conclusions. Second, when generalized to an overall population, our weighted model yielded odds ratios that were close, yet slightly higher, than those of our traditional bivariate model. This suggests that when adjusted to an entire population, the results were accurate, consistent, unbiased, and reliable to represent the overall population. Third, having at least one health condition or impairment influenced the overall communication difficulty with the doctor and significantly affected the responses to the four individual questions. Such difficulty answering these questions due to chronic health conditions leads to problems communicating with a doctor in the medical office. Thus, those with communication disorders must seek medical assistance, which can be in the form of a support aid or proxy.

## Conclusion

In conclusion, our longitudinal research indicates that participants with some hearing loss were the primary sensory factor contributing to difficulties in doctor-patient communication. In contrast, those with vision loss did not have the same impact. Minority status and low educational background were the most significant socioeconomic factors, while poor health conditions and challenges in daily tasks were the most critical medical factors. Future research must explore strategies to support individuals with these high-impact conditions and improve communication in healthcare settings for this disability group [15]. For instance, individuals with hearing disabilities could benefit from requesting accommodations such as assistive listening devices (e.g., audio induction loop systems) or utilizing interpreters, note takers, and captioning services. In addition to communication, future research should focus on improving physician relationships with patients who are often stigmatized as "disabled." Patients with chronic illnesses who struggle with daily tasks may experience communication challenges, highlighting the need for medical, social, and emotional support. Ongoing research and robust medical support from proxies and family members are crucial in addressing patient disparities and enhancing clinical outcomes.

## Acknowledgments section

National Health and Aging Trends Study. Produced and distributed by www.nhats.org with funding from the National Institute on Aging (grant number U01AG032947).

## Ethical approval and informed consent statements

The article consists of secondary data analysis of the National Health and Aging Trends Study (NHATS) Public Use Files. There are no human participants in this article and informed consent is not required.

## Data availability statement

The data used in our analysis is publicly available through a public use file on nhats.org, but users must first register for an account to access it.

